# Genomic epidemiology of SARS-CoV-2 in Mauritius reveals a new wave of infections dominated by the B.1.1.318, a variant under investigation

**DOI:** 10.1101/2021.06.16.21259017

**Authors:** Houriiyah Tegally, Magalutcheemee Ramuth, Daniel Amoaka, Cathrine Scheepers, Eduan Wilkinson, Marta Giovanetti, Richard J Lessells, Jennifer Giandhari, Arshad Ismail, Darren Martin, Emmanuel James San, Margaret Crawford, Rodney S Daniels, Ruth Harvey, Somduthsingh Bahadoor, Janaki Sonoo, Myriam Timol, Lovena Veerapa-Mangroo, Anne von Gottberg, Jinal N. Bhiman, Tulio de Oliveira, Shyam Manraj

## Abstract

Mauritius, a small island in the Indian Ocean, has had a unique experience of the SARS-CoV-2 pandemic. In March 2020, Mauritius endured a small first wave and quickly implemented control measures which allowed elimination of local transmission of SARS-CoV-2. When borders to the island reopened, it was accompanied by mandatory quarantine and testing of incoming passengers to avoid reintroduction of the virus into the community. As variants of concern (VOCs) emerged elsewhere in the world, Mauritius began using genomic surveillance to keep track of quarantined cases of these variants. In March 2021, another local outbreak occurred, and sequencing was used to investigate this new wave of local infections. Here, we analyze 154 SARS-CoV-2 viral genomes from Mauritius, which represent 12% of all the infections seem in Mauritius, these were both from specimens of incoming passengers before March 2021 and those of cases during the second wave. Our findings indicate that despite the presence of known VOCs Beta (B.1.351) and Alpha (B.1.1.7) among quarantined passengers, the second wave of local SARS-CoV-2 infections in Mauritius was caused by a single introduction and dominant circulation of the B.1.1.318 virus. The B.1.1.318 variant is characterized by fourteen non-synonymous mutations in the S-gene, with five encoded amino acid substitutions (T95I, E484K, D614G, P681H, D796H) and one deletion (Y144del) in the Spike glycoprotein. This variant seems to be increasing in prevalence and it is now present in 34 countries. This study highlights that despite having stopped the introduction of more transmissible VOCs by travel quarantines, a single undetected introduction of a B.1.1.318 lineage virus was enough to initiate a large local outbreak in Mauritius and demonstrated the need for continuous genomic surveillance to fully inform public health decisions.

## Introduction

Genomic surveillance has been an important tool for characterizing the COVID-19 pandemic, through mapping dissemination dynamics of the virus across the world or tracking the local movement and population dynamics of the virus within countries, cities and communities (1– 6). Consequently, by the end of May 2021, the GISAID database contained more than 1.6 million SARS-CoV-2 near-full genome sequences (7). In November and December 2020, after months of ongoing transmission in many areas of the world, genomic surveillance in the UK, South Africa, and Brazil detected three SARS-CoV-2 variants of concern (VOCs): Alpha (B.1.1.7) (8), Beta (501Y.V2/B.1.351) (9) and Gamma (P.1) (10). These VOCs each have multiple mutations, predominantly in the S-gene, that have subsequently been found to be associated with greater resistance to immunity induced by infection/vaccination, enhanced transmissibility, and/or increased pathogenicity (11–14).

As these VOCs began spreading throughout the world via air travel (15), the impact of travel restrictions and traveler quarantines were again debated. A few countries which had been successful at near-elimination of COVID-19, turned to the sequencing of variants within SARS-CoV-2 positive travelers to inform public health measures aimed at keeping VOCs at bay. Sydney, Australia, for example, first detected an outbreak of the Beta variant in a travel quarantine facility and quickly followed through with a city-wide lockdown for a few days (16).

Mauritius, a small island in the Indian Ocean with 1.2 million inhabitants, has had a unique experience of, and response to, the COVID-19 pandemic. Since early 2020, before the identification of the first cases in the country, Mauritius began requiring 14-day quarantine for incoming passengers arriving from the most affected countries of Asia and Europe (17). On 18 March 2020, the country recorded its first three positive cases in travelers who had not been required to quarantine (not arriving from countries defined to be most at risk), and the following day the country went into a strict national lockdown, consisting of the closure of all non-essential economic activities, educational institutions and a ban of entry into the country. The first wave of infections lasted for approximately one month with a total of around 340 local cases across the island. With zero ongoing community cases following this, the country lifted all local restrictions on 15 June 2020 but borders remained closed. Total travel bans were eased on 1 October 2020 but a strict government managed 14-day hotel quarantine and regular RT-PCR tests for any incoming passengers were imposed by the government to prevent new infections from entering the community. As VOC emerged, travel bans were again imposed on passengers coming directly from the UK and South Africa. Between 30 April 2020 to February 2021, 23,471 passengers arrived in Mauritius through this quarantine system. Of these, 281 tested positive for COVID-19 during the said quarantine period. From June 2020 to February 2021, no local COVID-19 cases were recorded in the country. This unfortunately changed in March 2021 when a cluster of local cases was detected followed by a second wave of more than 650 local cases. So far, this second wave has been controlled with rigorous testing of symptomatic individuals, their contacts a significant number of individuals within targeted areas as well as by the isolation of cases, contact tracing and testing the contacts of those in quarantine.

Here we describe the use of SARS-CoV-2 sequencing for the genomic surveillance of both community and travel-associated COVID-19 cases in Mauritius to help investigate this second wave and to inform the public health response to avert future outbreaks. We demonstrate how coupling quarantining of incoming travelers with SARS-CoV-2 testing has helped to prevent multiple introductions of two known VOCs into the country. Genomic surveillance also revealed that the second wave was dominated by a single introduction to the country of a potential variant of interest (VOI), B.1.1.318. This VOI has fourteen amino acid mutations in the Spike protein, including the E484K and P681H, which are also found in other VOCs.

## Results

### Travel quarantine control of VOCs

From October 2020 to February 2021, Mauritius re-opened its borders to commercial flights and allowed the entry of both citizens and tourists under a strict testing and (managed) quarantine protocol. All visitors to Mauritius were required to provide confirmation of a negative PCR test 48 hours prior to flight-boarding and had to quarantine for 14 days in a hotel, undergoing three more RT-PCR tests at days 0, 7 and 14 of the quarantine period, all at their own cost. During this time, the country recorded a low and stable number of positive cases in quarantine facilities (Fig 1A), from which infected individuals were immediately transferred to treatment centers and isolated until testing negative again. Sequencing of residual diagnostic samples from these individuals revealed multiple cases of infection with VOCs. There were six independent introductions (including two paired travelers) of the Beta (501Y.V2/B.1.351) variant from Africa and Europe, and two independent introductions of the Alpha (B.1.1.7) from Europe (Fig 1B, Supp Fig S1). Sequencing of specimens from these travelers also detected one case of the A.23.1 variant, three cases of the European lineage B.1.177.4 and an infection incident of one health worker in a quarantine facility by two travel-related cases carrying the B.1.36 lineage (Supp Figure S1). The B.1.36 lineage was common in India, where these travelers had come from at the beginning of 2021. Subsequent sequencing of community cases during the second wave of local infections in March 2021 (124 of 432 positive cases sequenced in March 2021 suggested that the VOCs detected in quarantine did not spill over into the community (Fig 1B).

**Figure 1.**
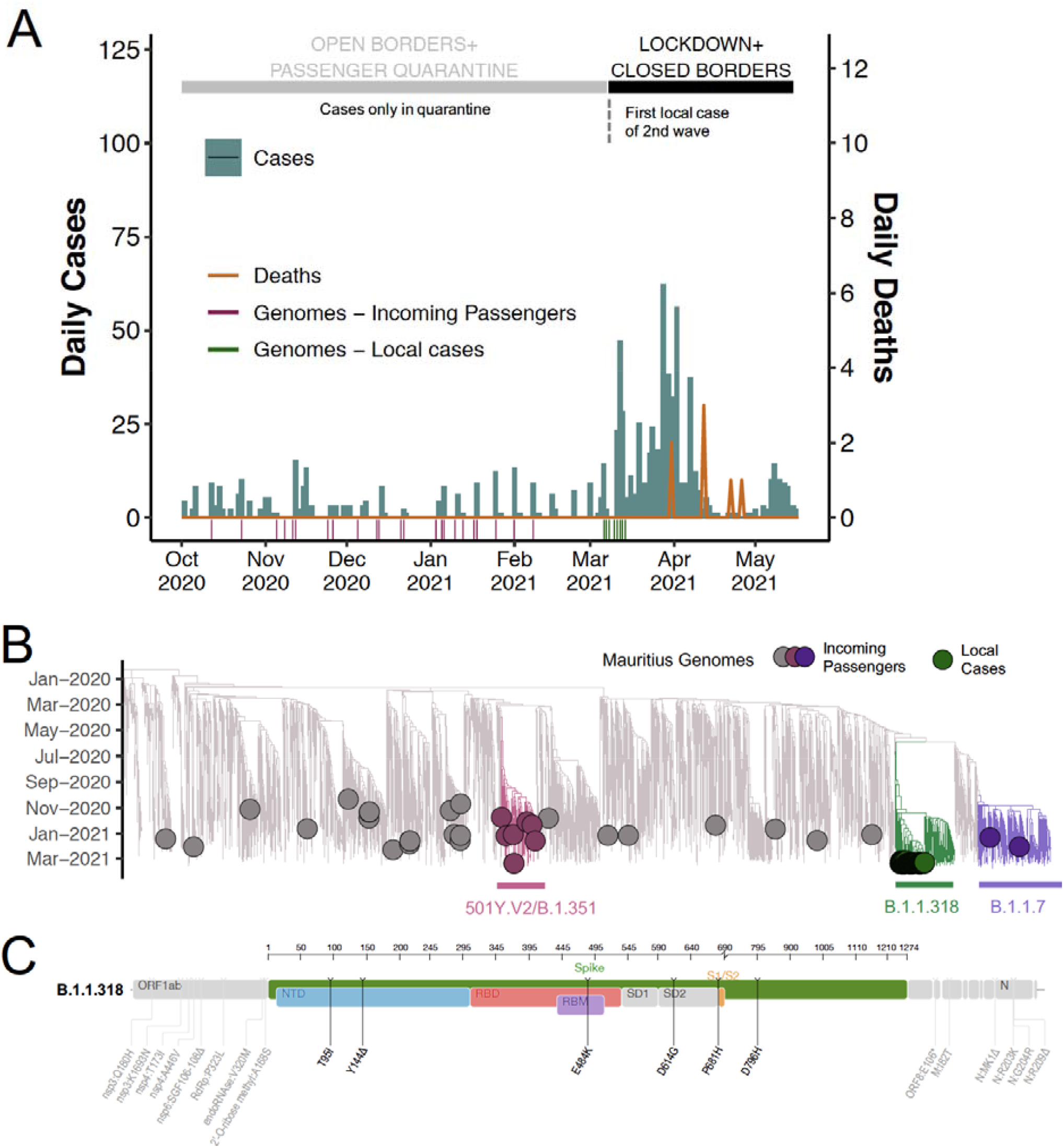
B.1.1.318 variants dominating second wave of infections in Mauritius. A) Epidemiological curve showing daily number of COVID-19 cases (blue) and associated deaths (orange) in Mauritius from October 2020 to May 2021, alongside a timeline of the status of lockdown and border control status. The dates of genome sampling are also shown, distinguishing genomes from incoming passengers (n=34) (pink) or local cases (n=120) (green). B) ML tree of 4484 genomes, including 154 genomes from Mauritius, colored to show VOCs Alpha (501Y.V2/B.1.351) and Beta (B.1.1.7) and the VOI B.1.1.318. C) Protein map of amino acid mutations in the B.1.1.318 VOI, with a zoomed focus on the Spike glycoprotein.

### Second wave dominated by B.1.1.318, potential VOI

On 5 March 2021, Mauritius detected a first community case of COVID-19 in what would become the country’s second wave of infections. Epidemiologically, the second wave was characterized by multiple clusters in family, workplace and health facility settings and the country went into its second lockdown on 9 March 2021. At the peak of the second wave, Mauritius recorded 68 cases in one day on 11 March 2021 with 2555 tests carried out that day (positivity rate of 2.6%) following an average of approximately 1500 tests per day during this second wave (Source: Government Information Services Mauritius). Unfortunately, the country also recorded seven new COVID-related deaths during this time. To understand the origins and dynamics of this new wave of infections and to assess the potential impact of the lineages on the planned vaccine roll-out, virus-positive samples from 120 cases from the March 2021 outbreak were sequenced. The geographical sampling of these sequenced cases closely matched the concentration of cases during this outbreak within the districts of Plaines Wilhems and Black River (Supp Fig S2 – genomes sampling map and cases map). These were in addition to the successful sequencing of 34 samples from quarantined incoming passengers, placing Mauritius in fifth position worldwide for the proportion of cases (n=1287) sequenced (12%) at this timepoint (Supp Table S1).

A maximum likelihood (ML) tree representing the evolutionary relationships of the 154 Mauritian genomes with a representative subset of 4330 genomes sampled globally indicated that all 120 of the genomes sampled from local community-derived infections fall within a cluster of sequences belonging to variant lineage B.1.1.318 (Fig 1B). This lineage is characterized by fourteen non-synonymous mutations in the S-gene, with five encoded amino acid substitutions (T95I, E484K, D614G, P681H, D796H) and one deletion (Y144del) in the Spike glycoprotein (Fig 1C). While this lineage has not been defined as a VOI by the World Health Organization (WHO), several of the Spike glycoprotein mutations are common to other VOCs and VOIs (Supp Fig S3), including mutations associated with immune escape (E484K, Y144del and likely T95I) and increased transmissibility (P681H). Moreover, all five of the spike codon sites where lineage-defining mutations have occurred are detectably evolving under positive selection (p <0.05 with both the MEME and IFEL positive selection detection methods) (Supp Table S2). Overall, therefore, phylogenetic analysis of this outbreak cluster in Mauritius suggests that it was initiated by a single undetected introduction of a B.1.1.318 variant virus.

### International epidemiology of the B.1.1.318 variant lineage

To contextualize the B.1.1.318 variant outbreak in Mauritius, we downloaded all sequences belonging to the B.1.1.318 lineage on GISAID as of 17 May 2021. We identified 34 countries in total where cases of infection with B.1.1.318 variants had been sequenced (Fig 2A). Although Mauritius is the only country where B.1.1.318 has accounted for the majority of infections (76% of all sequenced cases), this variant has been classified as a variant under investigation (VUI-202102/04) by Public Health England since February 2021 due to it carrying the Spike glycoprotein E484K substitution (18). While this variant has been consistently sampled since the beginning of 2021 in several countries, such as the UK, the US, Germany and Ireland, with first detection being in Nigeria (West Africa), it has not been shown to circulate elsewhere at such high proportions as was seen in Mauritius (Fig 2A). Other countries with the highest prevalence of B.1.1.318 variants are Gabon (accounting for 19% of all sequenced cases), Togo (7%), and Greece (3%) (Fig 2A).

**Figure 2.**
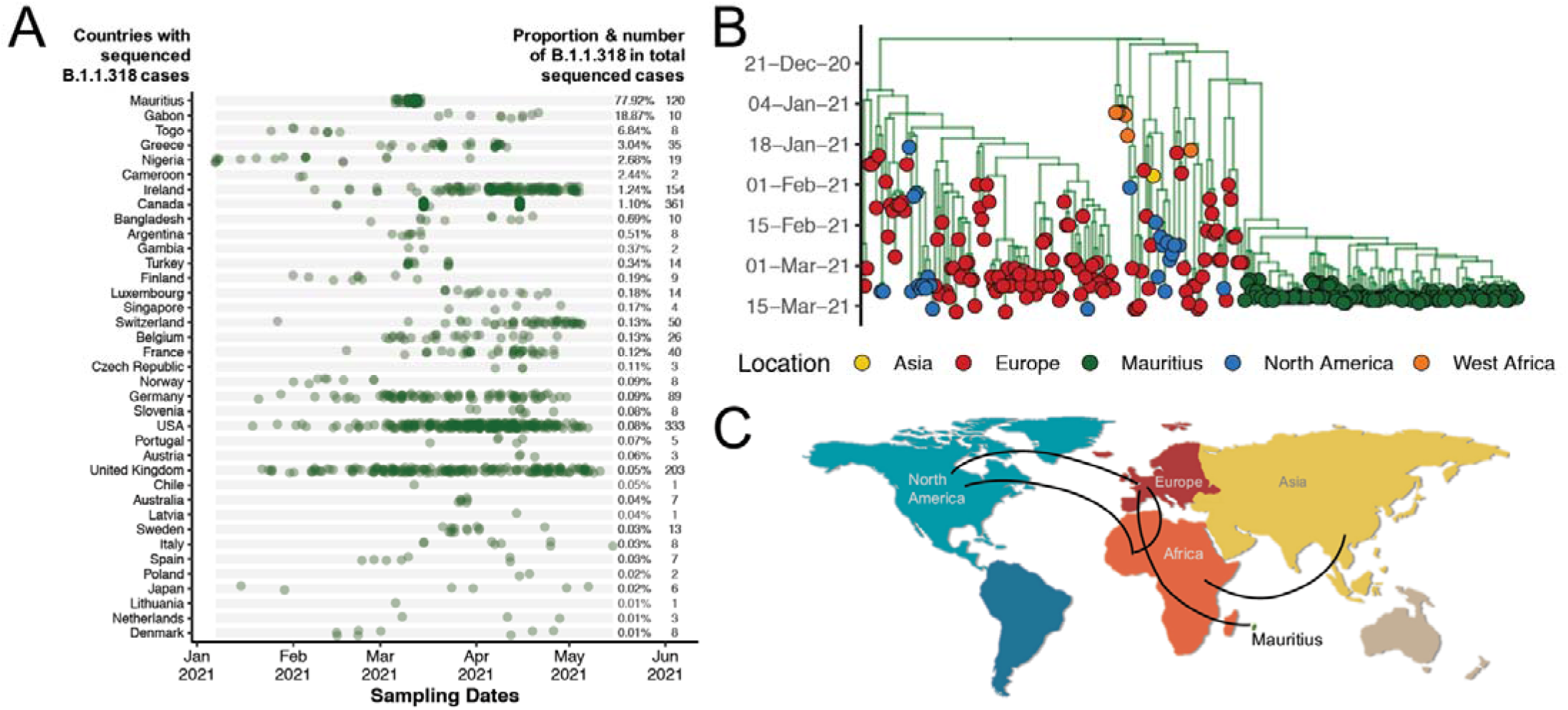
Global distribution and spread of the B.1.1.318 lineage. A) Prevalence of the B.1.1.318 lineage worldwide shown over time and as a proportion of all sequenced cases in each country. B) MCC tree of the B.1.1.318 lineage worldwide discretized by continental regions. C) International phylogeography of the B.1.1.318 lineage (lines showing spread in anti-clockwise direction).

A time-resolved Maximum Clade Credibility (MCC) analysis was performed with all B.1.1.318 variant sequences in the EpiCov™ database of GISAID. The resulting phylogeny indicates a potential origin of this lineage in Nigeria in late December 2020 (HPD: late December 2020 – early January 2021) (Fig 2B). The results show evidence of multiple introductions to Europe in January 2021 and onward transmission within Europe (Fig 2B). The lineage was introduced to North America both from Africa in late January 2021 and from Europe multiple times between mid-January 2021 and March 2021, with evidence of onward transmission in North America. Introduction of the lineage to Asia happened in late January 2021 with no evidence of onward transmission (Fig 2B, 2C). The introduction of the B.1.1.318 to Mauritius seems to have occurred via a single source around mid-February 2021 HPD: mid-February 2021 – late February 2021) (Fig 2B) and spread as small onward clusters within the country. The virus seems to have been imported to Mauritius from Europe with the closest relatives in the phylogeny being sequences of viruses from Ireland, Finland, and Norway (Fig 2C, Supp Fig S4).

## Discussion

Being a small island state, Mauritius was able to respond rapidly and aggressively to the first wave of SARS-CoV-2 infections in the country in March 2020. By closing borders and instituting an immediate strict lockdown, Mauritius was able to achieve zero transmission of the virus in the community and with ongoing public health and social measures, life resumed normally a couple of months later. In an attempt to revive tourism, borders reopened to both Mauritian citizens and foreigners in October 2020, but a strict testing and managed quarantine protocol was established to prevent any re-introduction of SARS-CoV-2 into the local community. As in Australia and New Zealand, this system proved to be effective in preventing new virus introductions and local outbreaks.

In fact, in this study we show that Mauritius stopped the introduction of more transmissible VOCs at least nine times due to this approach. For nine months, Mauritius was considered to be COVID-19 safe with minimal restrictions and COVID-19 protocols imposed on local residents. Unfortunately, the evidence presented here suggests that despite these effective border controls, a single undetected introduction of a B.1.1.318 lineage virus was enough to initiate a large local outbreak. It is as yet unknown how this variant was introduced to Mauritius but it is likely to have occurred through a porous layer of the quarantine system for travelers; sequencing of specimens from travelers from January to March 2021 is ongoing in an effort to trace a probable source. Given that there were no direct flights between Finland, Ireland and Norway to support the introduction of B.1.1.318 directly from these countries, it is plausible that the introduction may have occurred through intermediate links (such as travel hubs) and not be reflected in this genomic surveillance data. The current data demonstrates the dominance of B.1.1.318 infections in the second wave. However, ongoing sequencing of cases from later in the outbreak will rule out the possibility of the co-circulation of other variants.

While B.1.1.318 has been classified as a VUI by Public Health England (PHE), largely because of the presence of E484K, not much is known about this lineage other than it having three lineage-defining changes in the spike gene (Y144 Del, E484K and P681H) and one in nsp6 (106-108Del) that have convergently arisen in one or more of the better-known VOCs. All four of the defining amino acid substitutions in Spike (T95I, E484K, P681H and D796H) are at genome sites that have been detectably evolving under positive selection within the global SARS-CoV2 dataset since December 2020 when this lineage likely emerged. Further, the D796H mutation has been detected in a long-term SARS-CoV-2 infection, suggesting that it too might be significantly adaptive (19). Deletions within the NTD region of the Spike glycoprotein, especially in position 141-146, like the Y144Del seen in the B.1.1.318 virus, have been associated with resistance to several neutralizing antibodies but do not seem to confer significant reduction in vaccine efficacy (20). The P681H mutation, commonly present in the B.1.1.318 and the Alpha (B.1.1.7) variants, is proximal to the S1/S2 furin cleavage site of the Spike glycoprotein, a region known to impact transmissibility of the virus (12,21). Lastly, the E484K, found in the Receptor Binding Domain (RBD) of the Spike glycoprotein and also present in the B.1.1.318, the B.1.351, the P.1 and other variants, is known to cause escape to polyclonal antibodies and show decreased susceptibility both to convalescent and vaccine plasmas (14,22–24).

Although the B.1.1.318 has been detected in more than 30 countries on four continents and is currently still being detected, this lineage had not been able to outcompete VOCs in other regions of the world until recently when it became dominant in Greece. Although it seeded a large outbreak in Mauritius, it was introduced at a time when no other variants were locally transmitting, so it is unclear whether specific biological properties of B.1.1.318 viruses (such as increased transmissibility) may have contributed to initiation and propagation of the outbreak. Immune evasion properties introduced by Spike glycoprotein substitutions (e.g. E484K and Y144del) are unlikely to be relevant in explaining the outbreak as there was presumably a low level of population immunity following a small first wave. This may be relevant when weighing the importance of controlling the outbreak, to ensure the best chance of success of the local vaccination program. The vaccination campaign in Mauritius started in January 2021 for front-line workers and vulnerable individuals and in March 2021 was extended to the general adult population. Vaccines being used are Astrazeneca/Covishield, Covaxin and Sinopharm. At the end of May 2021, close to 20% of the population of Mauritius had received the first dose and half of those had also received the second dose of one of the vaccines, but vaccination is not yet thought to have played a major role in slowing down the second wave in March and April 2021.

In conclusion, SARS-CoV-2 genomic surveillance in Mauritius demonstrated a second wave that was dominated by a VUI, despite PCR testing and quarantine effectively preventing local transmission of other variants. This report also highlights the need for continuous genomic surveillance to fully to understand the transmission dynamics of the VUI in Mauritius.

## Data Availability

All of the genomes are available in GISAID.

## Acknowledgments

We are grateful to the Ministry of Health and Wellness of Mauritius for providing samples for this study. Equally, we are grateful to Mungra Priscilla, Mathur Hari, Sujeewon Chitra and all the staff of the Virology and Molecular Biology departments of the Victoria Hospital Mauritius for their dedication and hard work. We would also like to thank our colleagues Professor Aris Katzourakis from the University of Oxford and Dr. Gkikas Magiorkinis from National and Kapodistrian University of Athens for insightful discussions on the epidemic in Greece. We are immensely grateful to the laboratories and institutions around the world which have sampled and sequenced B.1.1.318 variants in their respective countries and made the data publicly available on GISAID (Supp Table S4), and to Public Health England for initially flagging this variant. These were all crucial in contextualizing our study. We also acknowledge Harshil Patel, Robert Goldstone, John Mccauley and Jerome Nicod at the Francis Crick Institute for assistance in sample management and data processing.

## Methods

### Epidemiological data

We analyzed COVID-19 cases and death counts in Mauritius from publicly released data up to 17 May 2021 from the Our World in Data (OWID) data repository (https://github.com/owid/covid-19-data). For correlation with government epidemic control measures, information was extracted from government press releases and speech transcripts.

### SARS-CoV-2 samples and metadata

We obtained deidentified remnant nasopharyngeal and oropharyngeal swab samples from patients testing positive for SARS-CoV-2 by RT-qPCR from the Ministry of Health and Wellness of Mauritius primary testing laboratory, the Central Health Laboratory at Victoria Hospital. These were used for SARS-CoV-2 Whole Genome Sequencing (WGS). RNA was extracted using the MagMAX™ Viral/Pathogen II Nucleic Acid Isolation Kit (Cat. No. A48383) using the automated nucleic acid extractor KingFisher™ Apex Magnetic Particle Processor with 96 Deep-Well Head using sample volume input of 200 µL. Associated metadata for the samples included date and location (district) of sampling, and sex and age of the patients.

### Real Time RT-PCR

In order to detect the SARS-CoV-2 virus by RT-PCR, the TaqPath COVID-19 CE-IVD RT-PCR Kit (Life Technologies, Carlsbad, CA) was used according to the manufacturer’s instructions. The assays target genomic regions (ORF1ab, S protein and N protein) of the SARS-CoV-2 genome. RT-PCR was performed on a QuantStudio 7 Flex Real-Time PCR instrument (Life Technologies, Carlsbad, CA). Cycle threshold (Ct) values were analyzed using auto-analysis settings with the threshold lines falling within the exponential phase of the fluorescence curves and above any background signal.

### WGS and genome assembly

RNA was extracted manually using the Qiagen Viral RNA Mini Kit (QIAGEN, California, USA). cDNA synthesis was performed on the RNA using random primers followed by gene specific multiplex PCR using the ARTIC protocol (25). Briefly, extracted RNA was converted to cDNA using the Superscript IV First Strand synthesis system (Life Technologies, Carlsbad, CA) and random hexamer primers. SARS-CoV-2 whole genome amplification by multiplex PCR was carried out using primers designed on Primal Scheme (http://primal.zibraproject.org/) to generate 400bp amplicons with an overlap of 70bp that covers the 30Kb SARS-CoV-2 genome. PCR products were cleaned up using AmpureXP purification beads (Beckman Coulter, High Wycombe, UK) and quantified using the Qubit dsDNA High Sensitivity assay on the Qubit 4.0 instrument (Life Technologies Carlsbad, CA).

The Illumina® Nextera Flex DNA Library Prep kit was used according to the manufacturer’s protocol to prepare uniquely indexed paired end libraries of genomic DNA. Sequencing libraries were normalized to 2nM, pooled and denatured with 0.2N sodium hydroxide. 1.3pM sample library was spiked with 1% PhiX (PhiX Control v3 adapter-ligated library used as a control). Libraries were loaded onto a 300-cycle NextSeq 500/550 Mid Output Kit v2. and run on the Illumina NextSeq550 instrument (Illumina, San Diego, CA, USA).

Raw reads coming from Illumina sequencing were assembled using Exatype (https://sars-cov-2.exatype.com/), Galaxy ARTIC pipeline (2. https://covid19.galaxyproject.org/artic/), Genome Detective 1.126 (https://www.genomedetective.com/) and the Coronavirus Typing Tool (26,27). The initial assembly obtained from Genome Detective was polished by aligning mapped reads to the references and filtering out low-quality mutations using bcftools 1.7-2 mpileup method.

Seventeen of the samples in this study were sequenced at the Francis Crick Institute on ONT GridION following the ARTIC LoCost V3 protocol (28). Data was demultiplexed and processed using v1.1.1 of the ncov2019-artic-nf pipeline [https://github.com/connor-lab/ncov2019-artic-nf] with the additional parameters “--schemeVersion V3 -- minReadsPerBarcode 1 --minReadsArticGuppyPlex 1”.

All of the sequences were deposited in GISAID (https://www.gisaid.org/) (29) and the GISAID accession included as part of the Supplementary Data (Table S3).

### Global reference dataset

Mauritian sequences were analyzed against a backdrop of globally representative SARS-CoV-2 genotypes. At the time of sequence analysis, more than 1.5 million SARS-CoV-2 genomes had been shared publicly. Due to the sheer size of this dataset and over sampling in specific countries, a systematic down-sampling was carried out. Important lineage defining genomes along with randomly sampled genotypes per location were included in the phylogenetic reconstruction. The dataset was also enriched with sequences from the B.1.1.318 lineage. The final reference set of 4484 genomes contained 635 from Africa, 582 from Asia, 1690 from Europe, 735 and 465 from North and South America respectively and 377 genotypes from Oceania.

### Phylogenetic analysis of SARS-CoV-2 in Mauritius

Mauritian sequences were analyzed against the global reference dataset using a custom build of the SARS-CoV-2 NextStrain pipeline (https://github.com/nextstrain/ncov) (30). The pipeline contains several python scripts that manage the analysis workflow. In short it allows for the filtering of genotypes, the alignment of genotypes in MAFFT (31), phylogenetic tree inference in IQ-Tree(32), tree dating and ancestral state construction and annotation. The resulting time scaled phylogeny can be viewed interactively and has been shared publicly on the NGS-SA NextStrain page (https://nextstrain.org/groups/ngs-sa/COVID19-Mauritius-2021.05.10). It was also visualized using R package ggtree (33).

### Lineage & Clade classification

We used the dynamic lineage classification method proposed in this study via the Phylogenetic Assignment of named Global Outbreak LINeages (PANGOLin) software suite (https://github.com/hCoV-2019/pangolin) (34). This was aimed at identifying the most epidemiologically important lineages of SARS-CoV-2 at the time of analysis, allowing researchers to monitor the epidemic in a specific geographical region. Accordingly, with this recently proposed dynamic lineage classification, many factors might suggest a new lineage including: i) monophyletic clusters on a global tree; ii) the presence of a statistically significant support (bootstrap/ultrafast bootstrap) on the node of the new lineages; iii) introduction into a novel geographic region; iv) epidemiological support (location; travel history) and v) characteristic Single Nucleotide Polymorphisms. Accordingly, three main SARS-CoV-2 lineages with those characteristics are currently recognized; lineage A, defined by Wuhan/WH04/2020, lineage B, defined by Wuhan-Hu-1 strain, and lineage C, a sub-classification from the B lineage. The BLAST function in GISAID (7) was also used to find sequences most similar to the Mauritius dataset and infer belonging to the B.1.1.318 lineage. We also classified the SARS-CoV-2 genomes in our dataset using the clade classification proposed by Nextstrain, divided into 19A, 19B, 20A, 20B, and 20C clades (35).

### Selection analysis

To identify which, if any, of the observed mutations in the Spike protein was most likely to increase viral fitness, we used the natural selection analysis of SARS-CoV-2 pipeline (https://observablehq.com/@spond/revised-sars-cov-2-analytics-page) (36,37). This pipeline examines the entire global SARS-CoV-2 nucleotide sequence dataset for evidence of:(*) (i) polymorphisms having arisen in multiple epidemiologically unlinked lineages, (ii) the likelihood of these polymorphisms being associated with a greater than expected ratio of non-synonymous:synonymous nucleotide substitution rates, and (iii) whether these polymorphisms have increased in frequency in the regions of the world where they have occurred.

### Dated phylogenetics

To estimate time-calibrated phylogenies dated from time-stamped genome data, we conducted phylogenetic analysis using the Bayesian software package BEASTv.1.10.4 (38), on a subset of data belonging to the B.1.1.318 lineage (n = 280) identified in the ML phylogeny to contain isolates from Mauritius(n=134). ML trees from this data subset was inspected in TempEst v1.5.3 (39) for the presence of a temporal (i.e. molecular clock) signal. Linear regression of root-to-tip genetic distances against sampling dates indicated that the SARS-CoV-2 sequences in this cluster evolve in a relatively-strong clock-like manner (r = 0.51; r^2^ = 0·3) (Supplementary Fig S4).

For this analysis we employed the strict molecular clock model, the HKY+I, nucleotide substitution model and the exponential growth coalescent model. We computed MCMC (Markov chain Monte Carlo) triplicate runs of 100 million states each, sampling every 10.000 steps for each data set. Convergence of MCMC chains was checked using Tracer v.1.7.141 (40). Maximum clade credibility trees were summarised from the MCMC samples using TreeAnnotator after discarding 10% as burn-in. We discretized sequence sampling locations as North America, Europe, Asia, West Africa and Mauritius.

### Phylogeographic analysis

To model phylogenetic diffusion of the B.1.1.318 cluster in Mauritius, we used a flexible relaxed random walk (RRW) diffusion model that accommodates branch-specific variation in rates of dispersal with a Cauchy distribution. For each sequence, latitude and longitude were attributed to a point randomly sampled within the patient’s district of residence.

MCMC chains were run for >100 million generations and sampled every 10000th step, with convergence assessed using Tracer v1.7 (40). Maximum clade credibility trees were summarized using TreeAnnotator after discarding 10% as burn-in. We used the R package “seraphim” to extract and map spatiotemporal information embedded in posterior trees (41).

## Supplementary Figures

**Supplementary Fig S1.**
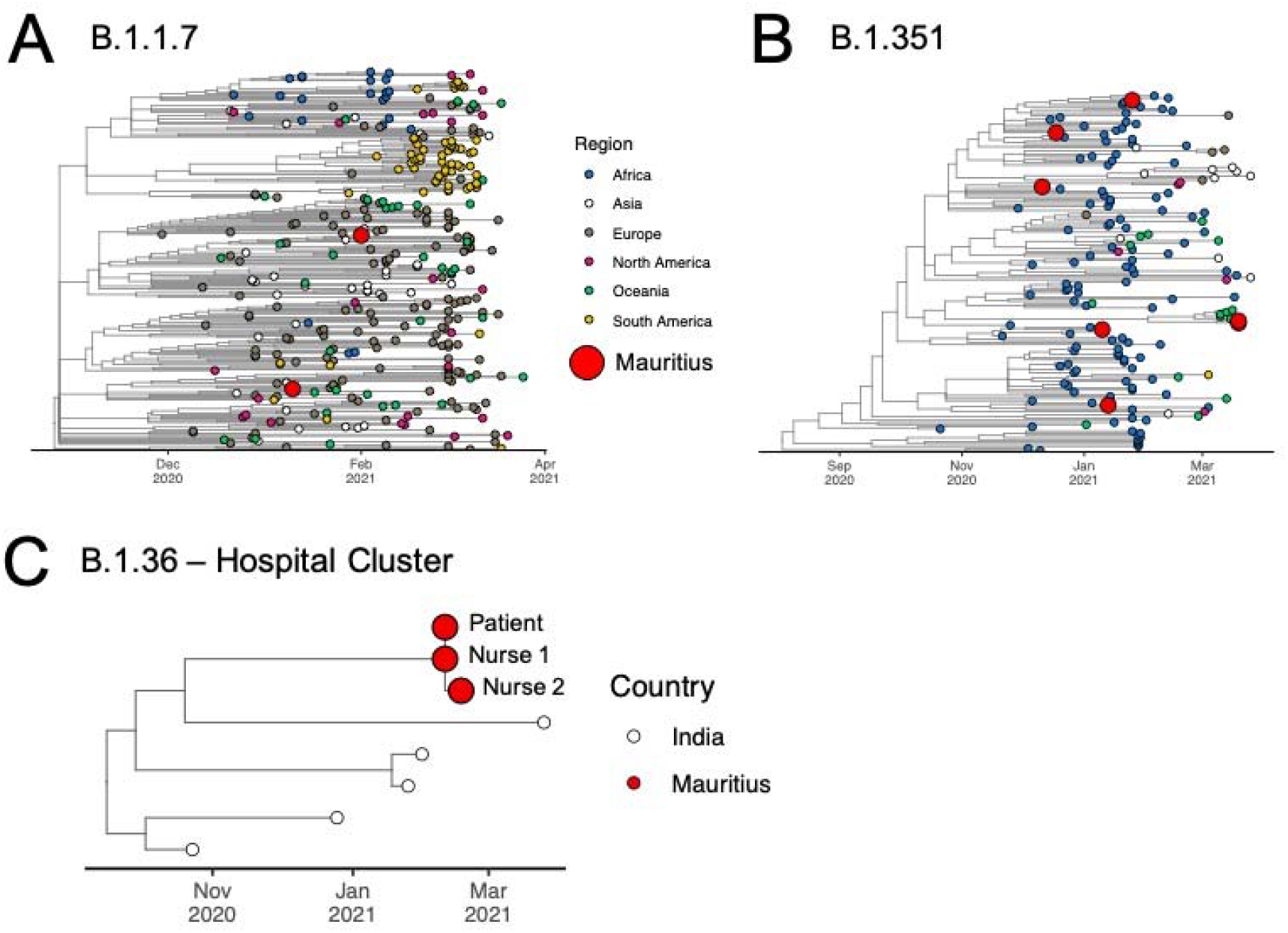
ML Timetree for three clusters of interest for introductions of SARS-CoV-2 to Mauritius (into quarantine centres): B.1.1.7 (A), 501Y.V2 (B), B.1.36 (C).

**Supplementary Fig S2.**
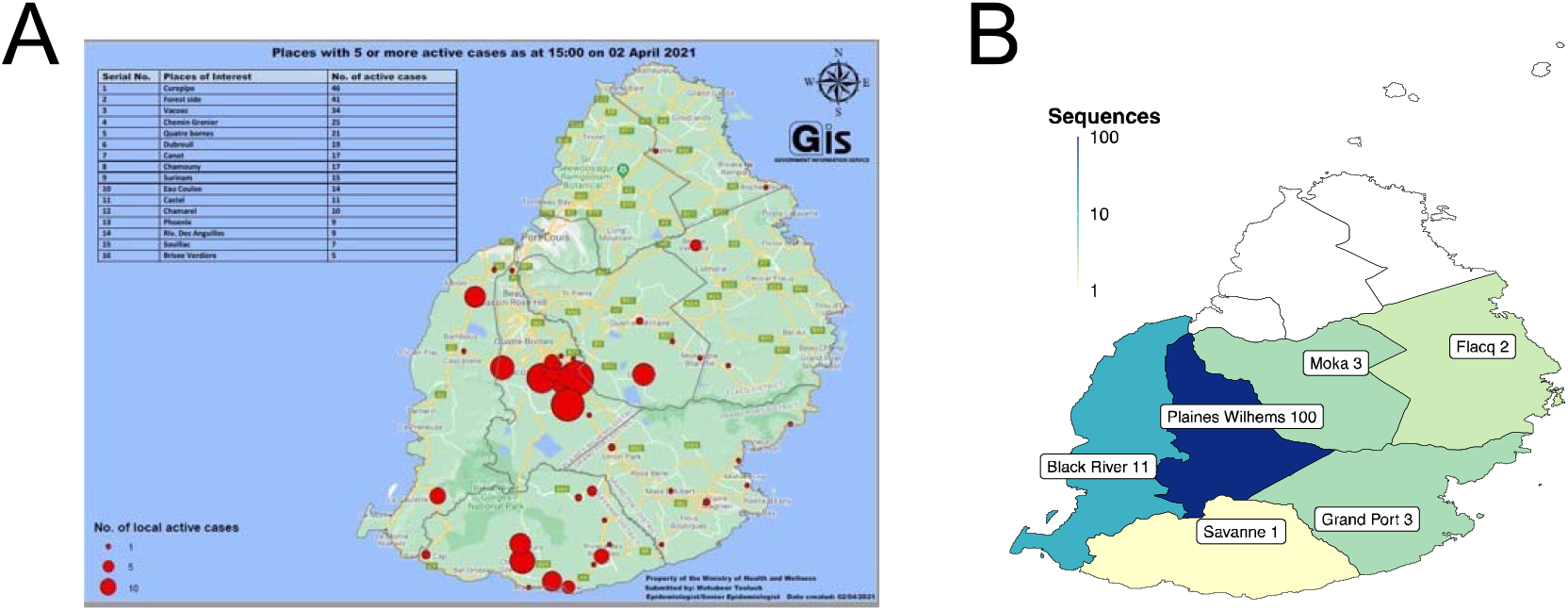
A) Geographical distribution of cases during the second wave in March 2021 (Picture property of Ministry of Health and Wellness of Mauritius and published by the Government Information Services on 2 April 2021) B) Location of sampling of genomes from the local (2^nd)^ second wave in March 2021 (n=120).

**Supplementary Fig S3.**
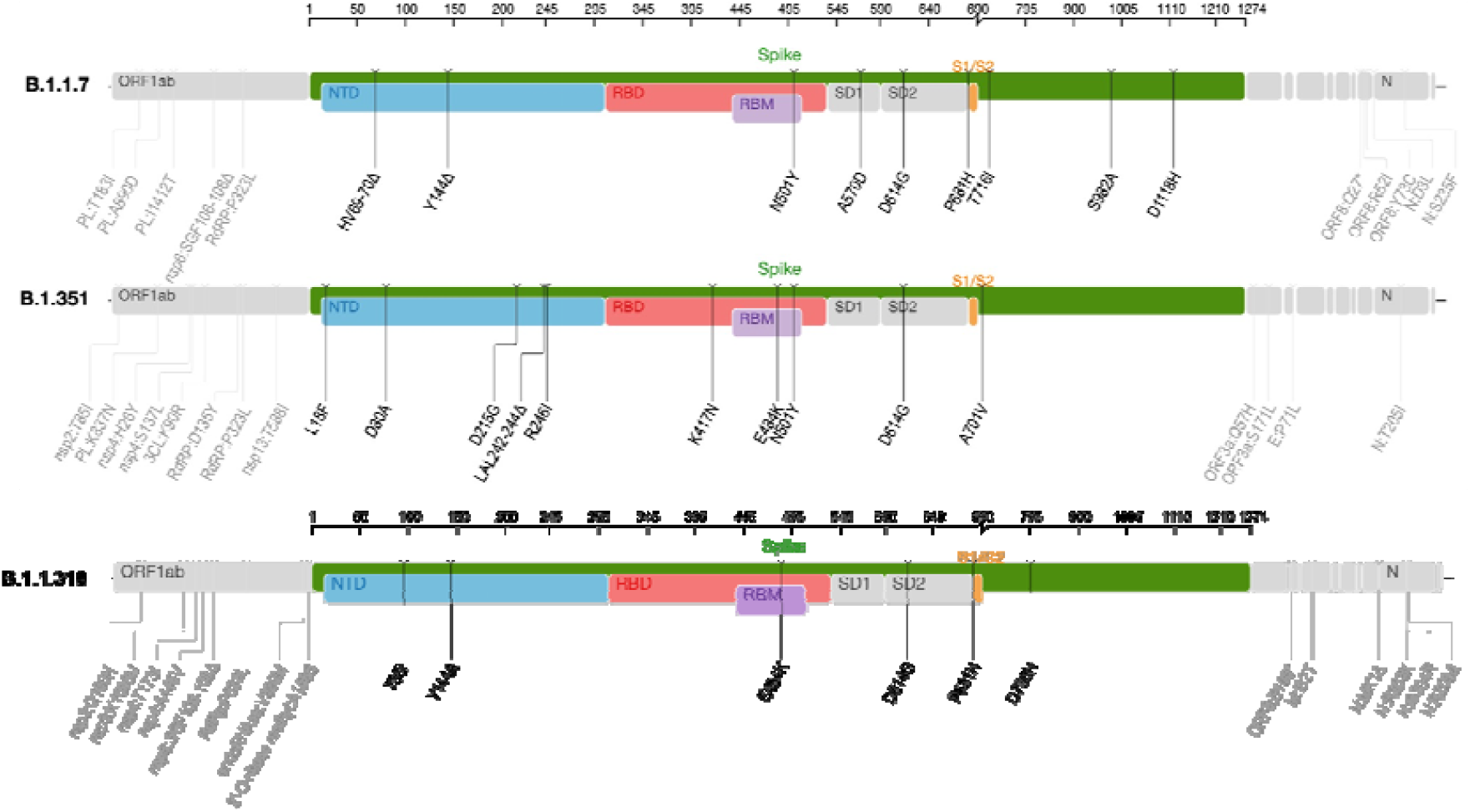
Genome maps showing a comparison of mutations between the B.1.1.318 variant and two known VOCs, the B.1.1.7 and the B.1.351

**Supplementary Fig S4.**
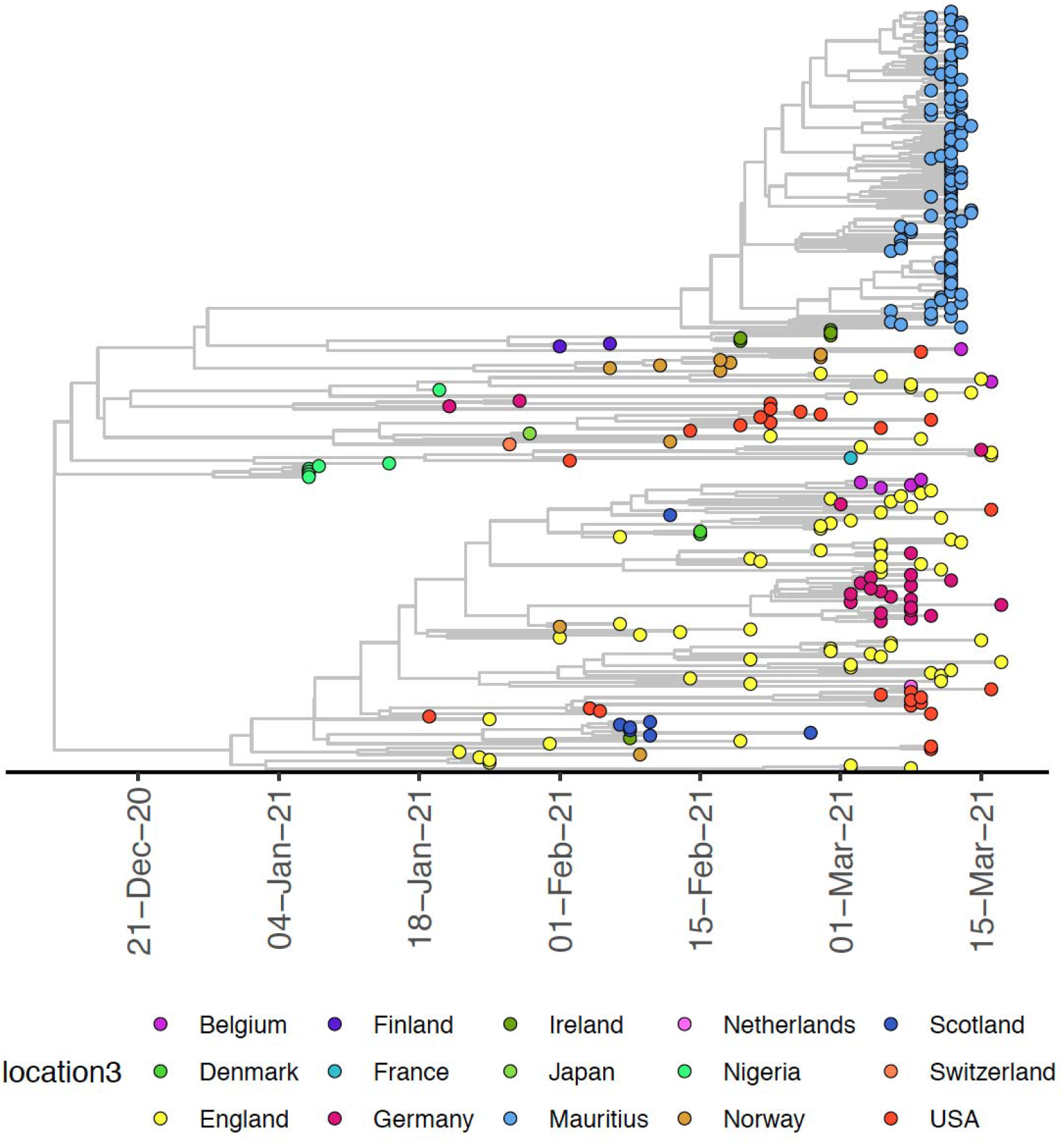
MCC tree of global B.1.1.318 sequences coloured by country

**Supplementary Fig S4.**
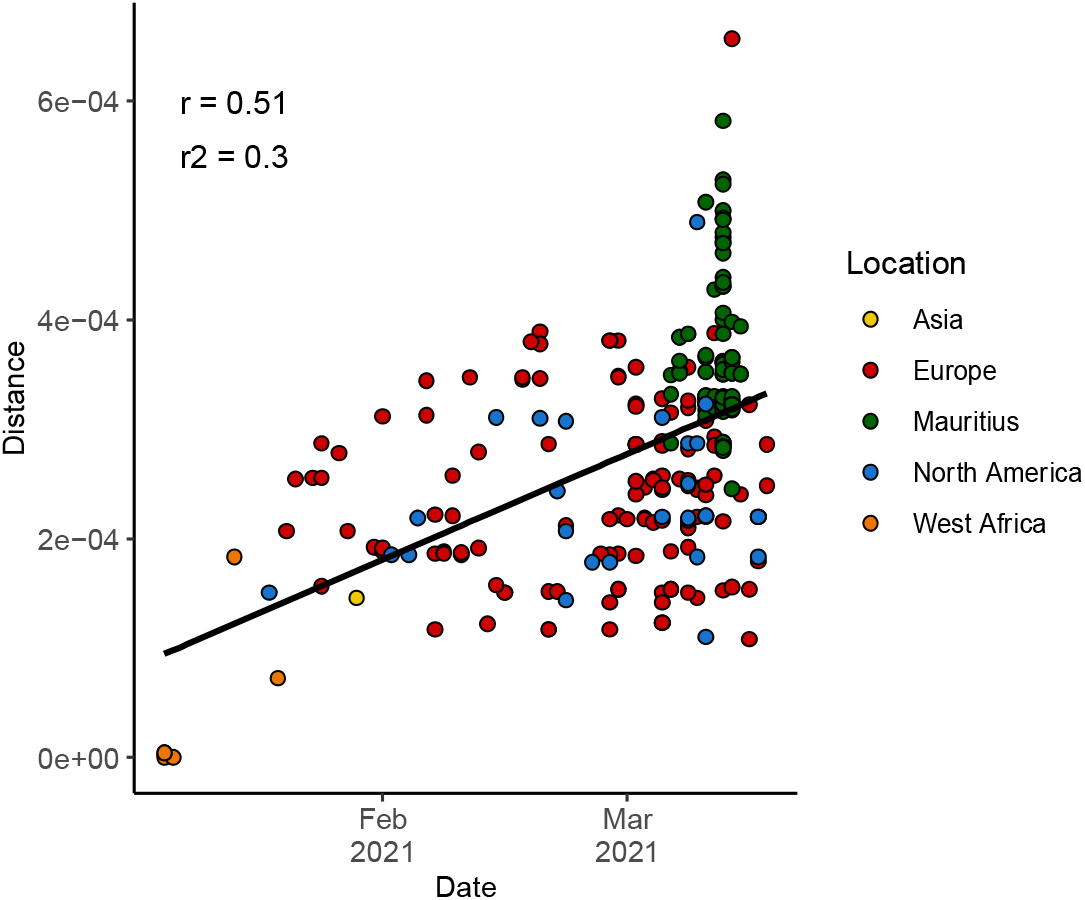
Root-to-tip regression plot of global B.1.1.318 lineage obtained through TempEst analysis showing the evolution of this lineage over time through accumulation of mutation (Distance) respective to the SARS-CoV-2 reference.

## Supplementary Tables

**Supplementary Table S1.** Ranking of the top 20 countries having sequenced the greatest proportions of SARS-CoV-2 cases relative to the total number of positive cases in the country.

| **Country** | **Number of Cases Sequenced** | **Total Cases (17 May 2021)** | **Percentage sequenced (%)** |
| --- | --- | --- | --- |
| Iceland | 5,085 | 6,548 | 77.7 |
| Australia | 18,174 | 29,983 | 60.6 |
| New Zealand | 1,282 | 2,653 | 48.3 |
| Denmark | 92,570 | 268,040 | 34.5 |
| Mauritius | 154 | 1,287 | 12.0 |
| Luxembourg | 7,771 | 69,190 | 11.2 |
| United Kingdom | 407,559 | 4,468,582 | 9.1 |
| Gambia | 537 | 5,957 | 9.0 |
| Taiwan | 179 | 2,017 | 8.9 |
| Norway | 8,668 | 119,500 | 7.3 |
| Saint Kitts and Nevis | 3 | 45 | 6.7 |
| Switzerland | 37,532 | 682,160 | 5.5 |
| Japan | 37,264 | 688,873 | 5.4 |
| Finland | 4,631 | 90,249 | 5.1 |
| Ireland | 12,397 | 254,870 | 4.9 |
| Brunei | 10 | 232 | 4.3 |
| Slovenia | 9,753 | 249,424 | 3.9 |
| Singapore | 2,406 | 61,613 | 3.9 |
| Sweden | 39,381 | 1,037,126 | 3.8 |
| Vietnam | 157 | 4,359 | 3.6 |

**Supplementary Table S2.**
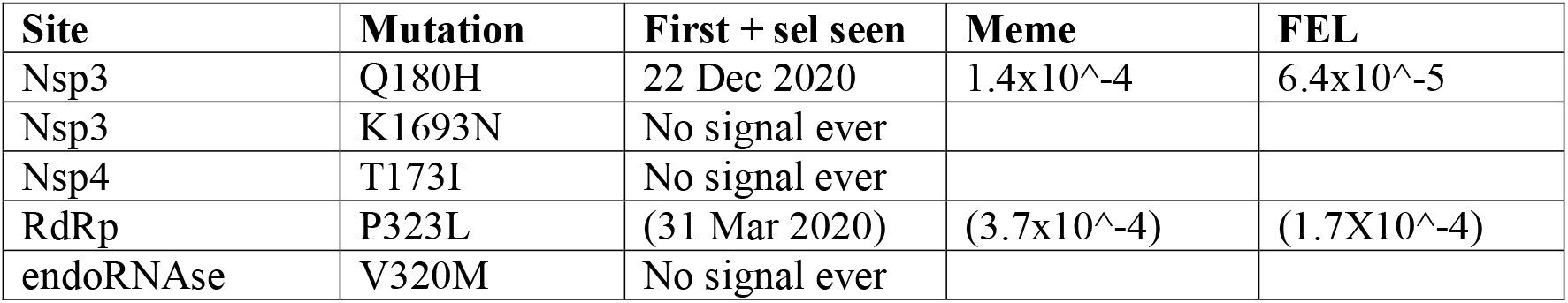
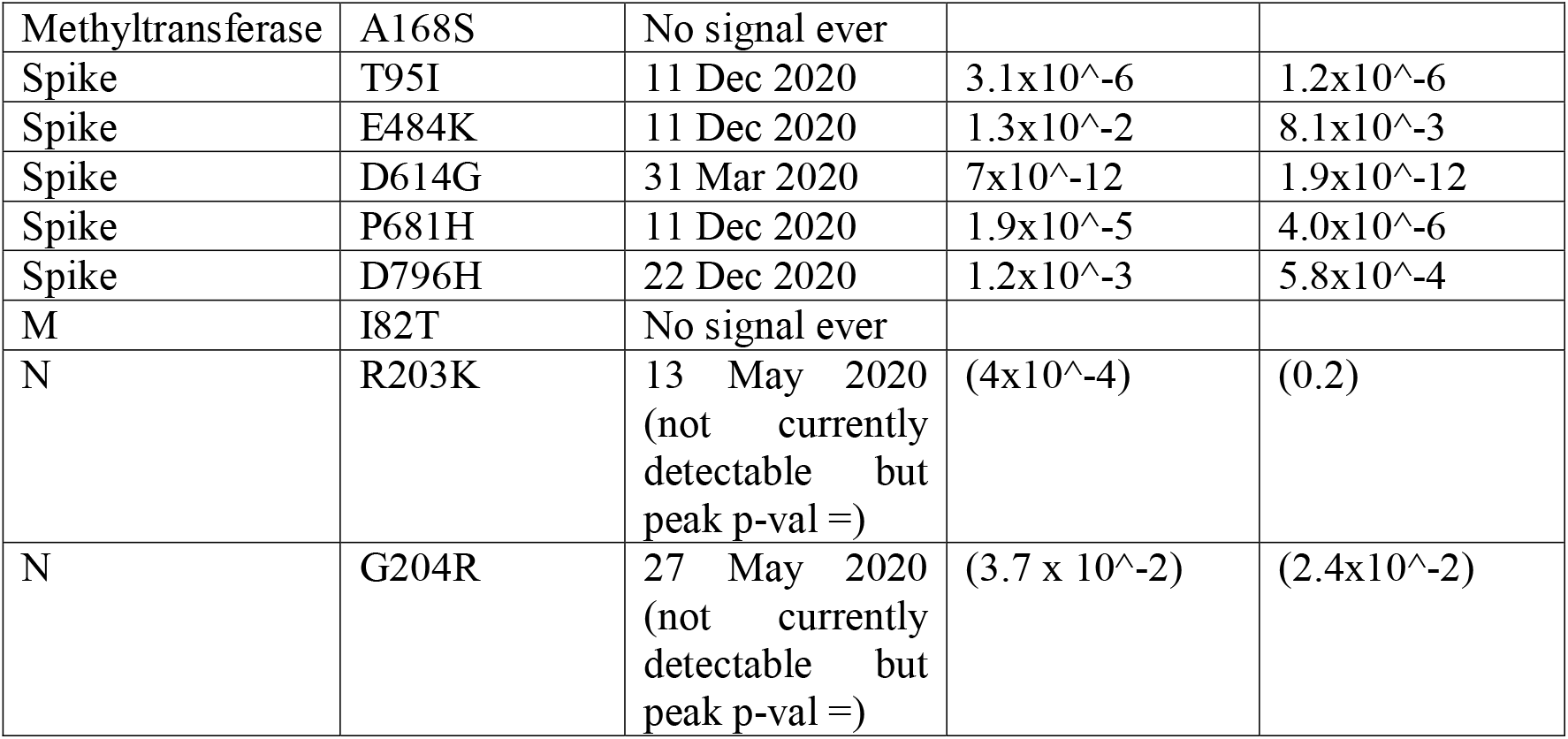
Sites of B.1.1.318 evolving under positive selection

**Supplementary Table S3.** Genomes from Mauritius on GISAID.

| strain | gisaid_epi_isl |
| --- | --- |
| hCoV-19/Mauritius/N2013/2020 | EPI_ISL_1191597 |
| hCoV-19/Mauritius/N2021/2021 | EPI_ISL_1191598 |
| hCoV-19/Mauritius/N2023/2021 | EPI_ISL_1191599 |
| hCoV-19/Mauritius/N2024/2021 | EPI_ISL_1191600 |
| hCoV-19/Mauritius/N2025/2021 | EPI_ISL_1191601 |
| hCoV-19/Mauritius/N2026/2021 | EPI_ISL_1191602 |
| hCoV-19/Mauritius/N2027/2021 | EPI_ISL_1191603 |
| hCoV-19/Mauritius/N2029/2021 | EPI_ISL_1191604 |
| hCoV-19/Mauritius/N2031/2021 | EPI_ISL_1191605 |
| hCoV-19/Mauritius/N2014/2020 | EPI_ISL_1191606 |
| hCoV-19/Mauritius/N2015/2020 | EPI_ISL_1191607 |
| hCoV-19/Mauritius/N2020/2021 | EPI_ISL_1191608 |
| hCoV-19/Mauritius/N4603/2021 | EPI_ISL_1827634 |
| hCoV-19/Mauritius/N4604/2021 | EPI_ISL_1827635 |
| hCoV-19/Mauritius/N4605/2021 | EPI_ISL_1827636 |
| hCoV-19/Mauritius/N4606/2021 | EPI_ISL_1827637 |
| hCoV-19/Mauritius/N4607/2021 | EPI_ISL_1827638 |
| hCoV-19/Mauritius/N4608/2021 | EPI_ISL_1827639 |
| hCoV-19/Mauritius/N4609/2021 | EPI_ISL_1827640 |
| hCoV-19/Mauritius/N4612/2021 | EPI_ISL_1827641 |
| hCoV-19/Mauritius/N4615/2021 | EPI_ISL_1827642 |
| hCoV-19/Mauritius/N4616/2021 | EPI_ISL_1827643 |
| hCoV-19/Mauritius/N4618/2021 | EPI_ISL_1827644 |
| hCoV-19/Mauritius/N4621/2021 | EPI_ISL_1827645 |
| hCoV-19/Mauritius/N4623/2021 | EPI_ISL_1827646 |
| hCoV-19/Mauritius/N4624/2021 | EPI_ISL_1827647 |
| hCoV-19/Mauritius/N4625/2021 | EPI_ISL_1827648 |
| hCoV-19/Mauritius/N4626/2021 | EPI_ISL_1827649 |
| hCoV-19/Mauritius/N4629/2021 | EPI_ISL_1827650 |
| hCoV-19/Mauritius/N4630/2021 | EPI_ISL_1827651 |
| hCoV-19/Mauritius/N4632/2021 | EPI_ISL_1827652 |
| hCoV-19/Mauritius/N4633/2021 | EPI_ISL_1827653 |
| hCoV-19/Mauritius/N4635/2021 | EPI_ISL_1827654 |
| hCoV-19/Mauritius/N4636/2021 | EPI_ISL_1827655 |
| hCoV-19/Mauritius/N4637/2021 | EPI_ISL_1827656 |
| hCoV-19/Mauritius/N4638/2021 | EPI_ISL_1827657 |
| hCoV-19/Mauritius/N4639/2021 | EPI_ISL_1827658 |
| hCoV-19/Mauritius/N4641/2021 | EPI_ISL_1827659 |
| hCoV-19/Mauritius/N4642/2021 | EPI_ISL_1827660 |
| hCoV-19/Mauritius/N4643/2021 | EPI_ISL_1827661 |
| hCoV-19/Mauritius/N4645/2021 | EPI_ISL_1827662 |
| hCoV-19/Mauritius/N4646/2021 | EPI_ISL_1827663 |
| hCoV-19/Mauritius/N4647/2021 | EPI_ISL_1827664 |
| hCoV-19/Mauritius/N4648/2021 | EPI_ISL_1827665 |
| hCoV-19/Mauritius/N4650/2021 | EPI_ISL_1827666 |
| hCoV-19/Mauritius/N4651/2021 | EPI_ISL_1827667 |
| hCoV-19/Mauritius/N4652/2021 | EPI_ISL_1827668 |
| hCoV-19/Mauritius/N4654/2021 | EPI_ISL_1827669 |
| hCoV-19/Mauritius/N4655/2021 | EPI_ISL_1827670 |
| hCoV-19/Mauritius/N4656/2021 | EPI_ISL_1827671 |
| hCoV-19/Mauritius/N4657/2021 | EPI_ISL_1827672 |
| hCoV-19/Mauritius/N4658/2021 | EPI_ISL_1827673 |
| hCoV-19/Mauritius/N4659/2021 | EPI_ISL_1827674 |
| hCoV-19/Mauritius/N4663/2021 | EPI_ISL_1827675 |
| hCoV-19/Mauritius/N4664/2021 | EPI_ISL_1827676 |
| hCoV-19/Mauritius/N4667/2021 | EPI_ISL_1827677 |
| hCoV-19/Mauritius/N4668/2021 | EPI_ISL_1827678 |
| hCoV-19/Mauritius/N4669/2021 | EPI_ISL_1827679 |
| hCoV-19/Mauritius/N4671/2021 | EPI_ISL_1827680 |
| hCoV-19/Mauritius/N4672/2021 | EPI_ISL_1827681 |
| hCoV-19/Mauritius/N4676/2021 | EPI_ISL_1827682 |
| hCoV-19/Mauritius/N4677/2021 | EPI_ISL_1827683 |
| hCoV-19/Mauritius/N4678/2021 | EPI_ISL_1827684 |
| hCoV-19/Mauritius/N4679/2021 | EPI_ISL_1827685 |
| hCoV-19/Mauritius/N4680/2021 | EPI_ISL_1827686 |
| hCoV-19/Mauritius/N4682/2021 | EPI_ISL_1827687 |
| hCoV-19/Mauritius/N4684/2021 | EPI_ISL_1827688 |
| hCoV-19/Mauritius/N4685/2021 | EPI_ISL_1827689 |
| hCoV-19/Mauritius/N4687/2021 | EPI_ISL_1827690 |
| hCoV-19/Mauritius/N4688/2021 | EPI_ISL_1827691 |
| hCoV-19/Mauritius/N4689/2021 | EPI_ISL_1827692 |
| hCoV-19/Mauritius/N4690/2021 | EPI_ISL_1827693 |
| hCoV-19/Mauritius/N4693/2021 | EPI_ISL_1827694 |
| hCoV-19/Mauritius/N4694/2021 | EPI_ISL_1827695 |
| hCoV-19/Mauritius/N4695/2021 | EPI_ISL_1827696 |
| hCoV-19/Mauritius/N4696/2021 | EPI_ISL_1827697 |
| hCoV-19/Mauritius/N4237/2021 | EPI_ISL_2285162 |
| hCoV-19/Mauritius/N4238/2021 | EPI_ISL_2285163 |
| hCoV-19/Mauritius/N4239/2021 | EPI_ISL_2285164 |
| hCoV-19/Mauritius/N4240/2021 | EPI_ISL_2285165 |
| hCoV-19/Mauritius/N4241/2021 | EPI_ISL_2285166 |
| hCoV-19/Mauritius/N4242/2021 | EPI_ISL_2285167 |
| hCoV-19/Mauritius/N4243/2021 | EPI_ISL_2285168 |
| hCoV-19/Mauritius/N4244/2021 | EPI_ISL_2285169 |
| hCoV-19/Mauritius/N4245/2021 | EPI_ISL_2285170 |
| hCoV-19/Mauritius/N4246/2021 | EPI_ISL_2285171 |
| hCoV-19/Mauritius/N4247/2021 | EPI_ISL_2285172 |
| hCoV-19/Mauritius/N4248/2021 | EPI_ISL_2285173 |
| hCoV-19/Mauritius/N4249/2021 | EPI_ISL_2285174 |
| hCoV-19/Mauritius/N4250/2021 | EPI_ISL_2285175 |
| hCoV-19/Mauritius/N4251/2021 | EPI_ISL_2285176 |
| hCoV-19/Mauritius/N4252/2021 | EPI_ISL_2285177 |
| hCoV-19/Mauritius/N4253/2021 | EPI_ISL_2285178 |
| hCoV-19/Mauritius/N4254/2021 | EPI_ISL_2285179 |
| hCoV-19/Mauritius/N4255/2021 | EPI_ISL_2285180 |
| hCoV-19/Mauritius/N4256/2021 | EPI_ISL_2285181 |
| hCoV-19/Mauritius/N4257/2021 | EPI_ISL_2285182 |
| hCoV-19/Mauritius/N4259/2021 | EPI_ISL_2285183 |
| hCoV-19/Mauritius/N4260/2021 | EPI_ISL_2285184 |
| hCoV-19/Mauritius/N4261/2021 | EPI_ISL_2285185 |
| hCoV-19/Mauritius/N4262/2021 | EPI_ISL_2285186 |
| hCoV-19/Mauritius/N4263/2021 | EPI_ISL_2285187 |
| hCoV-19/Mauritius/N4264/2021 | EPI_ISL_2285188 |
| hCoV-19/Mauritius/N4265/2021 | EPI_ISL_2285189 |
| hCoV-19/Mauritius/N4266/2021 | EPI_ISL_2285190 |
| hCoV-19/Mauritius/N4267/2021 | EPI_ISL_2285191 |
| hCoV-19/Mauritius/N4268/2021 | EPI_ISL_2285192 |
| hCoV-19/Mauritius/N4269/2021 | EPI_ISL_2285193 |
| hCoV-19/Mauritius/N4270/2021 | EPI_ISL_2285194 |
| hCoV-19/Mauritius/N4271/2021 | EPI_ISL_2285195 |
| hCoV-19/Mauritius/N4272/2021 | EPI_ISL_2285196 |
| hCoV-19/Mauritius/N4273/2021 | EPI_ISL_2285197 |
| hCoV-19/Mauritius/N4274/2021 | EPI_ISL_2285198 |
| hCoV-19/Mauritius/N4275/2021 | EPI_ISL_2285199 |

